# Neonatal asphyxia: associated factors among newborns delivered in Sao Tome & Principe

**DOI:** 10.1101/2025.11.09.25339852

**Authors:** Carolina Vasconcelos Baltazar, Marta Valério, Vitória Cadete, Beatriz Costa, Swasilanne Sousa, Nelson Bandeira, Filomena Pereira, Alexandra Vasconcelos

**Affiliations:** Department of Pediatrics, Hospital de Cascais Dr. José de Almeida, Lisboa, Portugal; Department of Pediatrics, ULS Lezíria, Santarém, Portugal; Department of Pediatrics, ULS S. José, Lisboa, Portugal; CEAUL - Centro de Estatística e Aplicações, Faculdade de Ciências, Universidade de Lisboa, Lisboa, Portugal; Department of Pediatrics, Hospital Dr. Ayres de Menezes, São Tomé, República Democrática de São Tomé e Príncipe; Department of Obstetrics and Gynecology, Hospital Dr. Ayres de Menezes, São Tomé, República Democrática de São Tomé e Príncipe; Unidade de Clínica Tropical - Global Health and Tropical Medicine (GHTM), Instituto de Higiene e Medicina Tropical (IHMT), Universidade NOVA de Lisboa, Lisboa, Portugal; Unidade de Clínica Tropical and Individual Health Care (IHC) - Global Health and Tropical Medicine (GHTM), Instituto de Higiene e Medicina Tropical (IHMT), Universidade NOVA de Lisboa, Lisboa, Portugal

## Abstract

**Background:** Neonatal asphyxia is a leading cause of neonatal morbidity and mortality, particularly in low- and middle-income countries where maternal and fetal healthcare resources are often limited. This study aims to identify the factors associated with neonatal asphyxia among newborns in Sao Tome and Principe, in order to improve clinical practices in maternal and perinatal care.

**Methods:** A hospital-based unmatched case-control study was conducted among newborns from randomly selected mothers. Cases were defined as newborns with a 5-minute APGAR score < 7, while controls had a 5-minute APGAR score between 7 and 10. Data were collected from antenatal care pregnancy cards, medical files and face-to-face interviews. Multivariable logistic regression was performed to identify independent risk factors for neonatal asphyxia, with statistical significance set at p < 0.05.

**Results:** Among 519 newborns included in the study, the prevalence of neonatal asphyxia was 8.1% (n = 42). Significant risk factors for neonatal asphyxia included maternal status as a student (aOR 5.30 95% CI 1.66─16.97; p=0.005), twin pregnancy (aOR 3.73 95% CI 1.21─11.47; p=0.022), mode of delivery other than normal vaginal deliver (aOR 46.24 95% CI 6.73─317.63; p<0.001), gestational age greater or equal to 41 weeks (aOR 4.57 95% CI 1.72─12.13; p=0.002), birth weight <2500g (aOR 5.34 95% CI 1.75─16.26; p=0.003) and infectious risk (aOR 11.07 95% CI 4.87─25.18; p<0.001). Cesarean section (aOR 0.02 95% CI 0.003─0.188; p<0.001) was found to be a protective factor.

**Conclusion:** Neonatal asphyxia was independently associated with maternal occupation as a student and multiple gestation (maternal factors); intrapartum infectious risk and dystocic delivery (intrapartum factors); as well as post-term gestation, and low birth weight (neonatal factors).

## Introduction

Neonatal asphyxia remains a major public health concern in perinatal medicine, particularly in low-resource settings [1]. It is defined as the newborn’s inability to initiate and sustain effective breathing at birth, resulting from impaired blood gas exchange that leads to hypoxemia and hypercapnia [2]. Immediate and effective resuscitation is essential to prevent significant neonatal morbidity and mortality [2, 3]. Without timely intervention, neonatal asphyxia can result in severe complications such as hypoxic-ischemic encephalopathy (HIE), which is associated with long-term neurodevelopmental impairment and substantial societal and economic burdens [3]. In low- and middle-income countries (LMICs), the burden of neonatal asphyxia is particularly high due to limited access to skilled birth attendants, inadequate resuscitation equipment, and delays in recognizing and managing perinatal complications [25–27].

The APGAR score—assessing appearance, pulse, grimace, activity, and respiration—remains a widely used and practical tool for the immediate evaluation of a newborn’s condition after birth [4]. Although the APGAR score alone has recognized limitations in accurately diagnosing neonatal asphyxia, especially in distinguishing its severity or underlying causes, it remains the most feasible and accessible method in resource-constrained settings where advanced diagnostic tools such as blood gas analysis or neuroimaging are often unavailable [3, 4]. The World Health Organization and other global health organizations continue to recommend the APGAR score as a key component of newborn assessment in LMICs [1, 3].

Numerous studies conducted in low- and middle-income countries (LMICs) have identified a wide range of contributing factors for birth asphyxia, which can be broadly categorized into antepartum, intrapartum, postpartum, and newborn-related risk factors [15–17]. Among these, intrapartum and newborn risk factors have consistently been recognized as the primary determinants of neonatal asphyxia, particularly in low-resource settings where access to timely and high-quality obstetric and neonatal care is often limited [20–21].

Intrapartum risk factors—such as prolonged or obstructed labor, fetal distress, meconium-stained amniotic fluid, and inadequate monitoring during labor—are frequently reported as leading causes of asphyxia in LMICs, due to delays in recognizing and managing complications [15, 18]. Newborn risk factors, including low birth weight, prematurity, and infections, also play a significant role, especially in settings where neonatal resuscitation and supportive care resources are scarce [16, 19].

The Democratic Republic of São Tomé and Príncipe (STP) is a sub-Saharan African country with a population of approximately 200,000, of whom 41% are under the age of 20 [5]. The country has made notable progress in reducing under-five mortality, with rates declining from 36 per 1,000 live births in 2010 to 14.5 in 2022 [6]. However, neonatal mortality remains a significant challenge, accounting for 7 deaths per 1,000 live births [6]. According to the World Health Organization, intrapartum complications—including birth asphyxia and birth trauma—are estimated to contribute to 26% of neonatal deaths in São Tomé and Príncipe [7]. This is consistent with broader trends observed in sub-Saharan Africa, where intrapartum-related events are among the leading causes of neonatal mortality [23–26].

Furthermore, a previous cohort study conducted by the authors [8–14] demonstrated that newborns with birth asphyxia had a 55-fold increased risk of perinatal and neonatal mortality [14]. These findings underscore the urgent need for improved perinatal care, timely identification of at-risk pregnancies, and the implementation of effective resuscitation protocols—particularly in low-resource settings where the impact of neonatal asphyxia is most profound [13–14, 27].

In São Tomé and Príncipe, as in many other low-resource settings, the burden of neonatal asphyxia remains high, in part due to systemic challenges such as shortages of skilled healthcare workers, limited access to essential equipment, and gaps in the continuum of care from pregnancy through delivery and the immediate postnatal period [21, 22].

This study aims to identify the factors associated with neonatal asphyxia among newborns in São Tomé and Príncipe, with the goal of informing and improving clinical practices in maternal and perinatal care. By understanding the most relevant risk factors in this context, targeted interventions can be developed to reduce the incidence of neonatal asphyxia and improve neonatal outcomes.

## Materials and methods

### Study design and period

A facility-based unmatched case–control study was conducted in STP among 519 newborns whose mothers gave birth at Hospital Dr. Ayres de Menezes (HAM) Maternity Unit. The recruitment of newborns’ mothers occurred from July 2016 to November 2018.

### Setting

Hospital Dr. Ayres de Menezes (HAM) is the sole tertiary healthcare facility on the island of Sao Tome, serving as the primary referral center for the most complex obstetric and neonatal cases from across the country. Approximately 98% of deliveries in Sao Tome occur in health facilities, with 82.4% taking place at the HAM maternity unit. HAM is the only center on the island equipped to provide Comprehensive Emergency Obstetric Care (CEmOC), including the capacity for blood transfusions and cesarean sections.

The hospital’s Neonatal Care Unit (NCU) has the capacity to admit up to six neonates and is able to manage uncomplicated neonatal conditions. However, the unit faces significant resource constraints, lacking critical interventions such as non-invasive and invasive ventilation, surfactant replacement therapy, electroencephalogram (EEG) monitoring, and parenteral nutrition. Furthermore, there are no neonatology specialists in the country; neonatal care is delivered by general practitioners and nursing staff, which may limit the ability to provide advanced neonatal care and manage severe complications. These constraints highlight the challenges faced in delivering high-quality neonatal care in low-resource settings and underscore the need for capacity building and resource allocation to improve neonatal outcomes in São Tomé and Príncipe.

### Participants

The eligibility criteria for study participants were: (1) all neonates delivered at Hospital Ayres de Menezes (HAM), and (2) newborns delivered outside HAM but admitted to the hospital on the day of birth. A total of 535 newborns were initially enrolled.

Exclusion criteria were as follows: (1) neonates delivered at HAM with no signs of life (stillbirths), (2) newborns whose mothers had cognitive impairment, and (3) adolescent or illiterate mothers who had not obtained consent from their parents or legal guardians to participate in the study. Sixteen newborns were excluded due to stillbirth, resulting in a final sample size of 519 participants.

### Selection of cases and controls

Cases were the asphyxia group (AG) were newborns with an APGAR-score at 5-minutes inferior to seven [4, 24]. Controls were the no asphyxia group (no AG) were newborns with an APGAR-score at 5-minutes between seven and ten.

### Sample size determination and sampling procedures

Sample size followed the WHO-steps approach [28] applying a web-based sample size calculator, Raosoft, which suggested a minimum sample size of S = 355, which placed the right dimension between 355 (95%) and 579 (99%) confidence [29]. A total of 535 participants were enrolled based on the following assumptions: two-sided 95% confidence level, and power of 80% to detect an odds ratio of at least 2 for neonatal asphyxia. This sample size was also supported by PASS software [30].

A random sampling was applied to recruit the newborn’s mother, selecting every second interval folder from the pile of mothers ’ medical folders, and occurred during daytime hours of working days. The mothers who consented were interviewed after recovery from the delivery, and the mother-newborn dyads were followed-up throughout their stays until hospital discharge.

### Data collection tools, procedures, and quality control

Data were collected using a pre-tested, structured, interviewer-administered questionnaire developed based on the Sao Tome and Principe Demographic and Health Survey (DHS) and relevant literature (23-27, 32). In addition, a standardized abstraction checklist was used to extract information from antenatal care (ANC) pregnancy cards and maternal and newborn medical records. The questionnaire captured data on parental socio-demographic characteristics, preconception healthcare utilization, and obstetric history.

To ensure data quality, the questionnaire was pretested on 10 cases and 13 controls, with subsequent modifications made to improve cultural appropriateness and clarity of terminology. All questionnaires were checked for completeness and consistency, and data collection procedures were regularly reviewed by supervisors. The principal investigator, a pediatrician, was responsible for all field activities, including: (i) obtaining informed consent and enrolling mothers, (ii) extracting data from ANC cards and medical records, (iii) performing clinical examinations of newborns to confirm diagnoses, (iv) conducting face-to-face interviews, and (v) entering all collected data into a digital survey tool. Double data entry was performed to minimize errors and ensure data accuracy.

### Operational definition of variables

**Table 1:** Operational definition of variables.

| Sociodemographic factors |  |
| --- | --- |
| **Residence** | urban residence if living in the capital city (Água Grande) and rural areas if living in all other districts (Cantagalo, Caué, Lobata, Lembá, Mé-Zochi and Príncipe island). |
| Preconception factors |  |
| **Poor birth spacing** | birth intervals of less than 2 years [33]. |
| Antenatal care factors |  |
| **Maternal anemia** | hemoglobin concentration <11 g/dL. |
| **Hyperglycemia** | serum glucose > 105 mg/dL. |
| Intrapartum factors |  |
| **Fetal malpresentation** | presenting fetal part other than cephalic (e.g breech, transverse, oblique) [34]. |
| **Prolonged rupture of membrane (PROM)** | rupture of membrane lasting > 18 hours before labor began [35, 36]. |
| **Pre-eclampsia** | hypertension ≥ 140/90 mmHg and proteinuria in dipsticks in women who were normotensive at antenatal care. |
| **Obstructed labor** | Sum of all cesarean sections due to mechanical problems or fetal distress and all instrumental delivery [37]. |
| **Postpartum hemorrhage** | bleeding > 500mL. |
| **Instrumental vaginal delivery** | Vacuum-assisted delivery (the only method used in STP). |
| Newborn factors |  |
| **Gestational age** | estimated from the date of onset of the last normal menstrual period or through ultrasound dating of pregnancy [38, 39]. Prematurity was defined as a delivery before 37 completed weeks of gestation [40], term when the delivery occurred between 37 and 40 weeks of gestation and post-term when occurred after 41 weeks of gestation. |
| **Infectious risk** | presence of (1) maternal fever (axillary temperature >37.9 C) at the time of delivery, (2) prolonged rupture of membrane (≥18 h), and/or (3) foul-smelling amniotic fluid [41]. |
| **Intrauterine growth restriction (IUGR)** | There is still no consensual definition for intrauterine growth restriction (IUGR) or fetal growth restriction in Africa [42]. |
| **Fetal distress at birth** | APGAR score < 7 at the first minute of life [24]. |

#### Data analysis

Data were entered into the QuickTapSurvey app (2010–2021 Formstack), an offline survey app tool, and exported to Excel to be further checked for completeness, coded, entered, and cleaned. Data were analyzed using SPSS version 25. In this study, cases were coded as 1, and controls were coded as 0 for analysis. The proportion of missing data ranged from 0.8 to 10% across variables. Missing values higher than 10% were described in the analysis.

Descriptive statistics were used to describe the frequency distribution of each of the variables mentioned earlier. The chi-square test was used to compare the proportion of cases and controls between selected categorical variables. To identify factors associated with neonatal asphyxia, univariable and multivariable logistic regression analyses were performed. In the univariable analysis, explanatory variables with a p value ≤0.25 were candidates for a multivariable logistic regression model to monitor the influence of confounding variables. With their 95% confidence intervals (95% CI), crude (cOR) and adjusted (aOR) odds ratios were determined to assess the strength and existence of an association. The level of significance α = 0.05 was considered.

#### Ethics approval and consent to participate

The study complies with the Declaration of Helsinki and was approved and consented to by dedicated ethics oversight bodies such as the Ministry of Health of STP and by the main board of HAM, since at the time the study protocol was submitted, there was no ethics committee in STP. All methods in our study were performed in accordance with the relevant guidelines and regulations in practice. Written informed consent was obtained from all participants after the purpose of the research was explained orally by the researcher. Approval by the participants ‘parents or legal guardians was asked in the case of adolescents under 16 years of age or for illiterate women. The participants or their legal representatives also consented to have the results of this research work published. Participation in the survey was voluntary, as participants could decline to participate at any time during the study.

## Results

A total of 519 newborns (42 cases and 477 controls) were enrolled in the study. The newborn’s mean gestational age (GA) was 38.73 weeks with a standard deviation (SD) of 2.62 weeks (minimum 22 weeks – maximum 43 weeks). The mean birth weight was 3053.79 g (± SD = 649 g) (minimum 900 g – maximum 4650 g). The current study revealed that 477 (91.9%) births had no neonatal asphyxia, while the remaining 42 (8.1%) were births with neonatal asphyxia. Cases had a mean GA and birth weight of 36.93 weeks (± SD = 4.74 weeks) and 2440.00 g (± SD = 921.94 g), respectively, while their counterparts had 38.89 weeks (± SD = 2.28 weeks) and 3107.83 g (± SD = 590.84 g), respectively, with significant difference (p<0.001). The newborn characteristics and complications are described in Table 2. The two groups showed significant differences in neonatal complications such as infectious risk (69% AG vs 18.4% no AG, p < 0.001), need for neonatal resuscitation (59.5% AG vs 0.6% nAG, p < 0.001), fetal distress at birth (100% AG vs 12,8% nAG, p < 0.001), admission at NCU (69% AG vs 8.6% nAG, p < 0.001) and administration of antibiotic therapy (73.8% AG vs 17% nAG, p < 0.001). The newborns with asphyxia tended to have more neonatal complications, which is consistent with previous research.

**Table 2:**
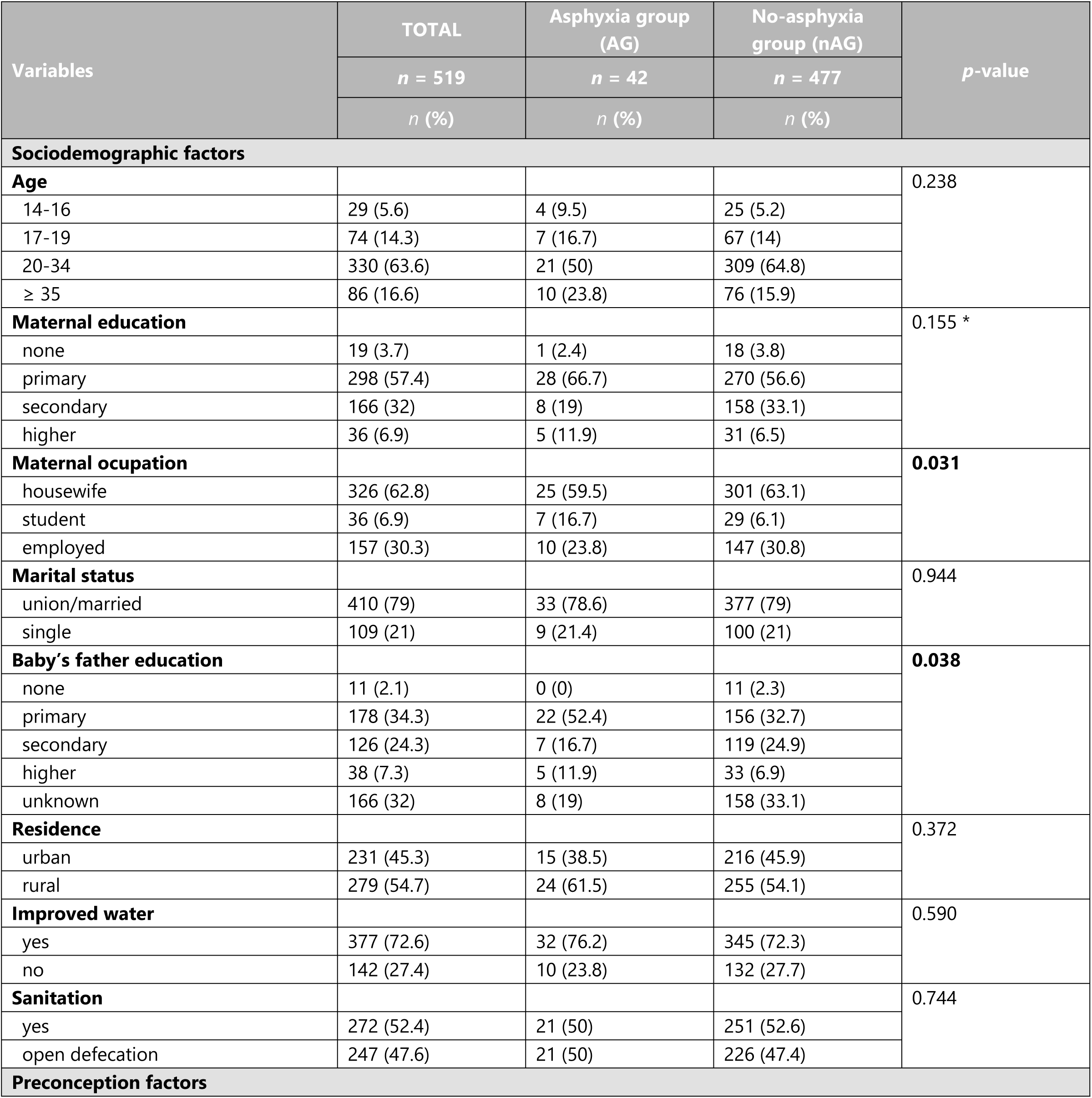
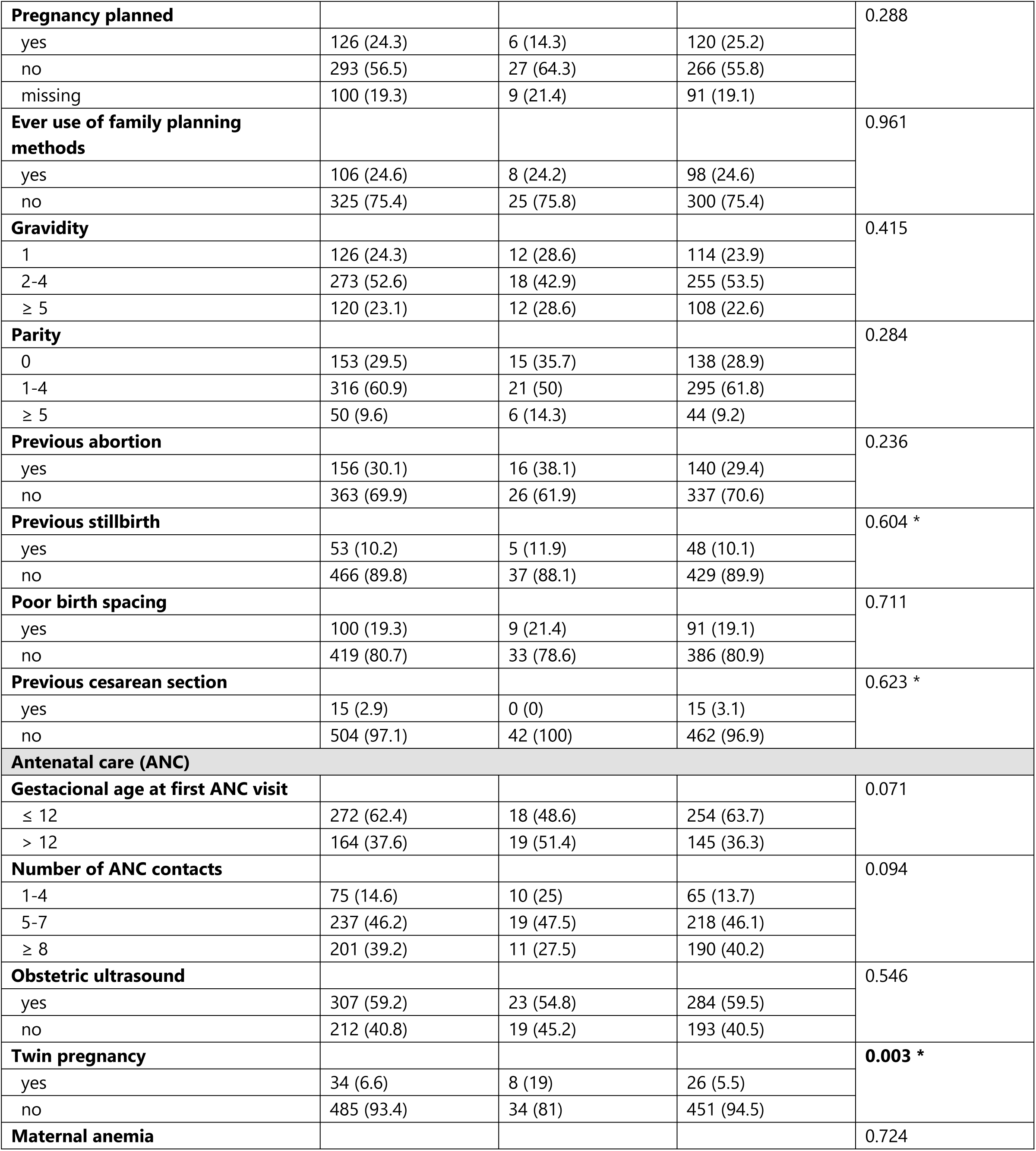
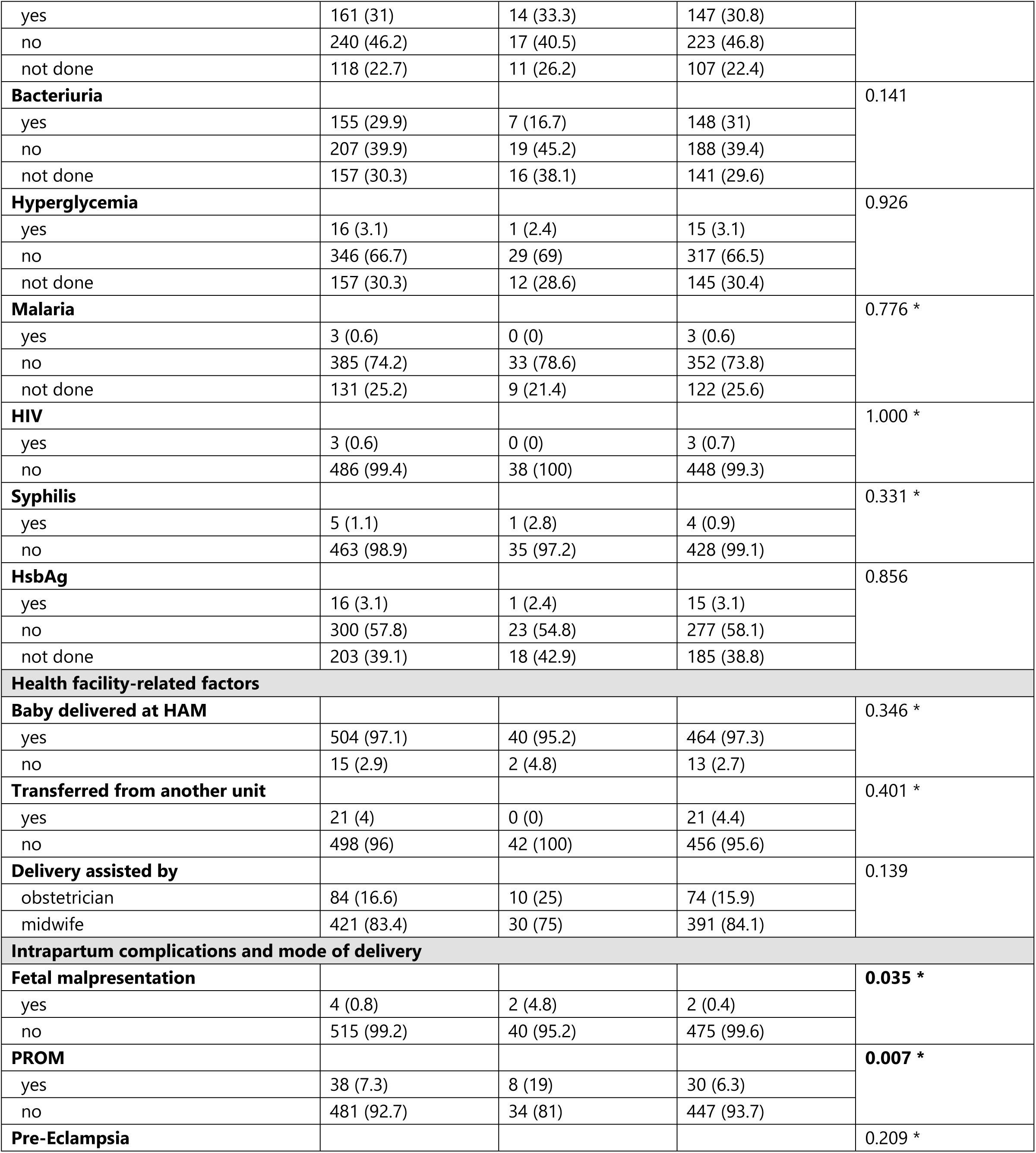
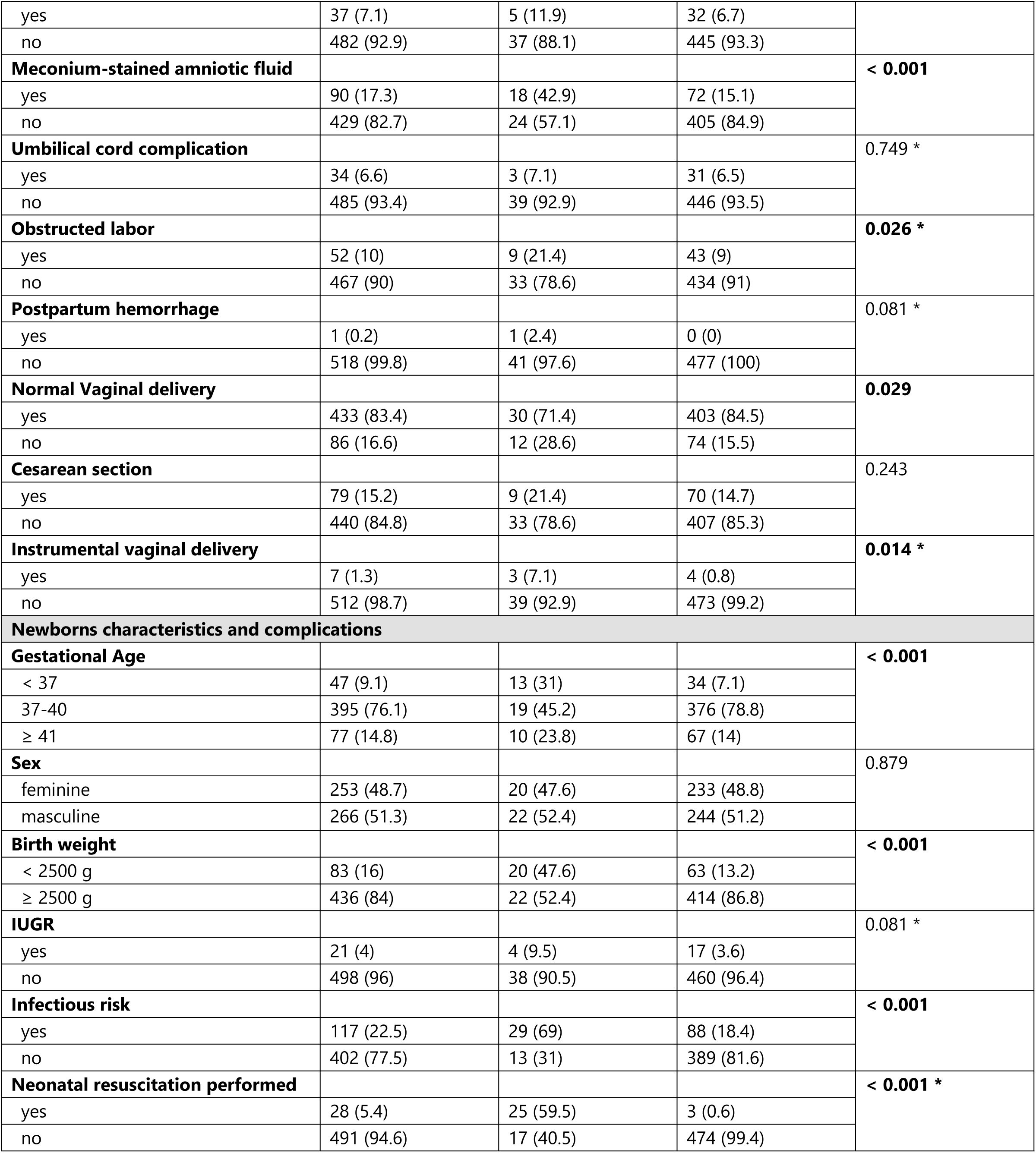
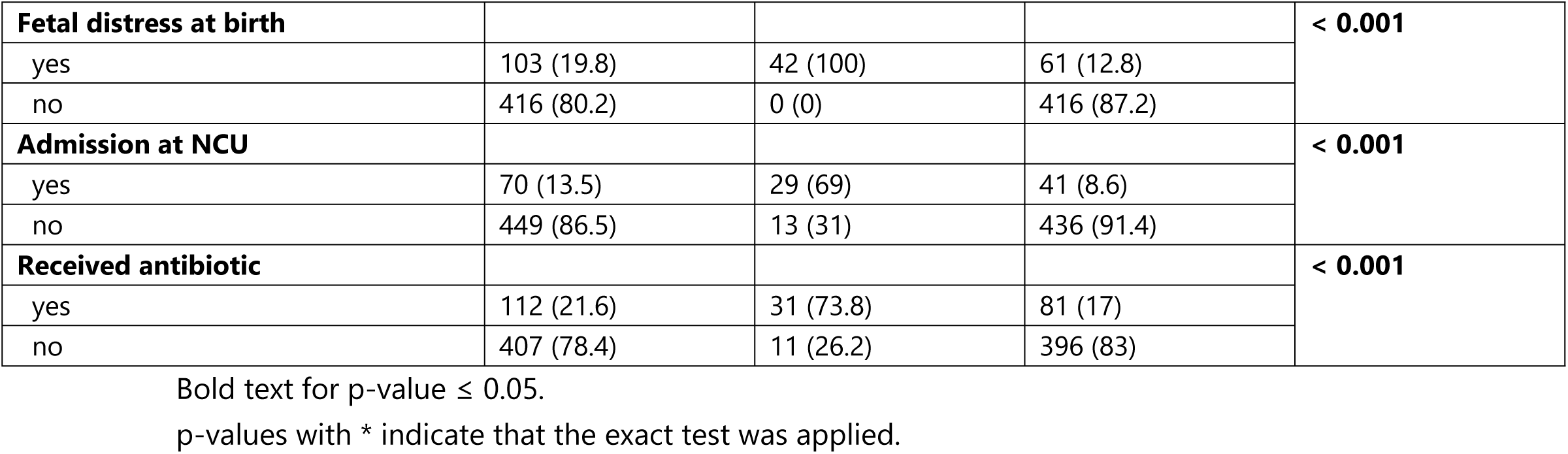
Maternal characteristics, antepartum, intrapartum and postpartum factors for all participants and for cases (newborns with asphyxia) versus controls (newborns without asphyxia) - frequencies, percentages and p-value (chi-Square test to compare proportions of cases and controls)

The mean maternal age was 26.5 years with a standard deviation of 7.03 years (minimum 14 years – maximum 43 years). The mean maternal age for cases and controls was 26.7 years (± SD = 7.83 years) and 26.5 years (± SD = 6.96 years), respectively. Maternal age did not differ significantly between groups (p = 0.238). The maternal characteristics as well as antepartum, intrapartum, and postpartum factors for all participants and for cases versus controls are described in Table 2. Concerning sociodemographic factors, only maternal occupation (p=0.031) and baby’s father education (p=0.038) showed significant differences between groups. The preconception factors studied (pregnancy planned, planning methods, gravidity, parity, previous abortion, previous stillbirth, poor birth spacing and previous cesarean section) did not differ significantly between the two groups. As for antenatal care, only twin pregnancy showed significant differences (19% AG vs 5.5% nAG, p=0.003). Health facility-related such as local of delivery, transfer for another unit and delivery assistance did not differ significantly between the asphyxia group and its counterparts. Relating the intrapartum complications and mode of delivery, fetal malpresentation (4.8% AG vs 0.4% nAG, p=0.035), PROM (19% AG vs 6.3% nAG, p=0.007), meconium-stained amniotic fluid (42.9% AG vs 15.1% nAG, p<0.001), obstructed labor (21.9% AG vs 9% nAG, p=0.026), vaginal delivery (71.4% AG vs 84.5% nAG, p=0.029) and instrumental vaginal delivery (7.1% AG vs 0.8% nAG, p=0.014) showed significant statistical differences.

### Factors associated with neonatal asphyxia

In the univariable logistic regression, maternal age, maternal occupation, pregnancy planned, parity, previous abortion, gestational age at first visit, number of ANC contacts, twin pregnancy, maternal bacteriuria, prolonged rupture of membranes, pre-eclampsia, meconium-stained amniotic fluid, obstructed labor, delivery assisted by, normal vaginal delivery, cesarean section, gestational age, birth weight, infectious risk were eligible for multivariable analysis (Table 3). Admission to the NCU and received antibiotic therapy were excluded from the multivariable analysis, as they represent consequences of adverse outcomes.

**Table 3:**
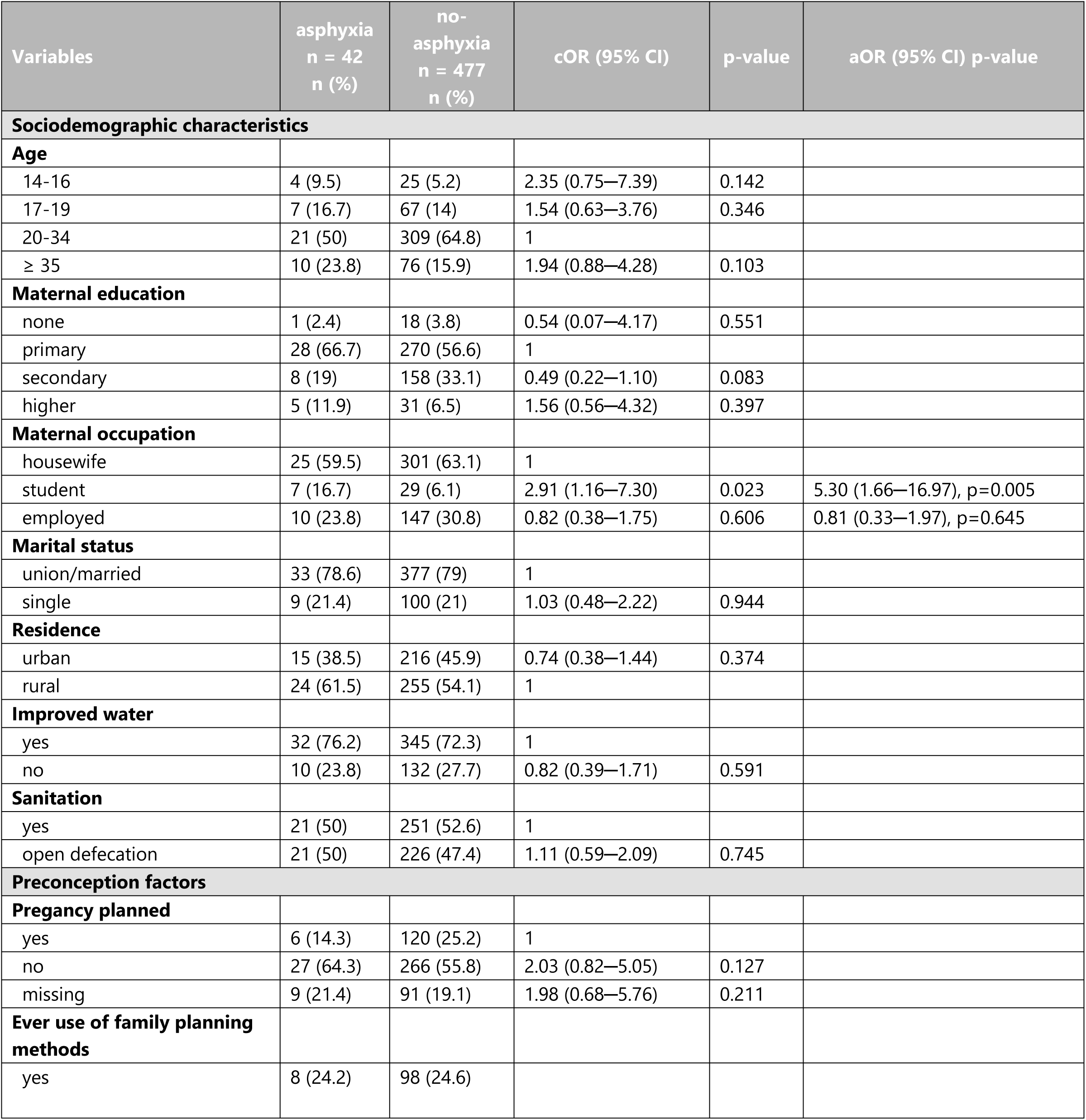
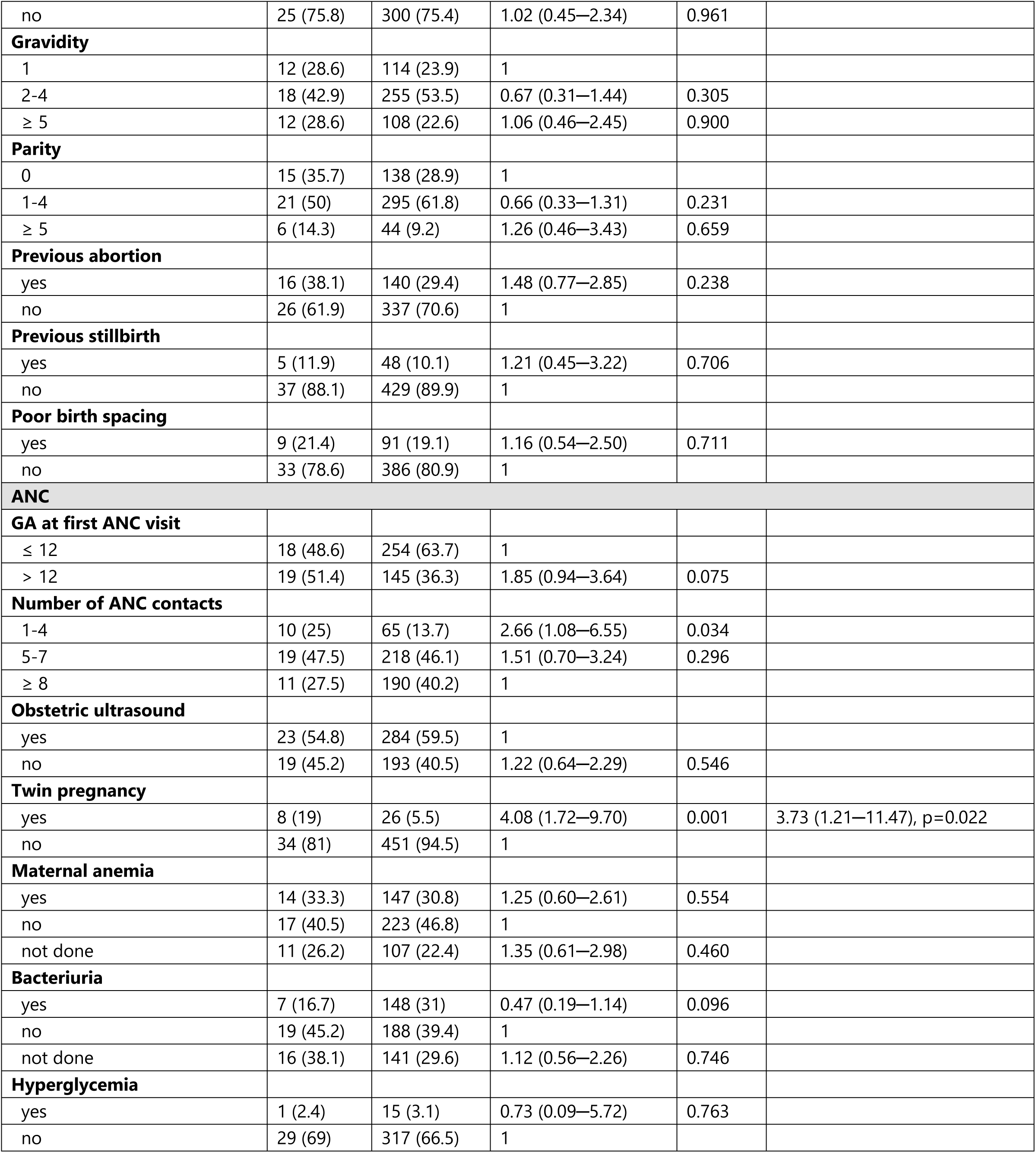
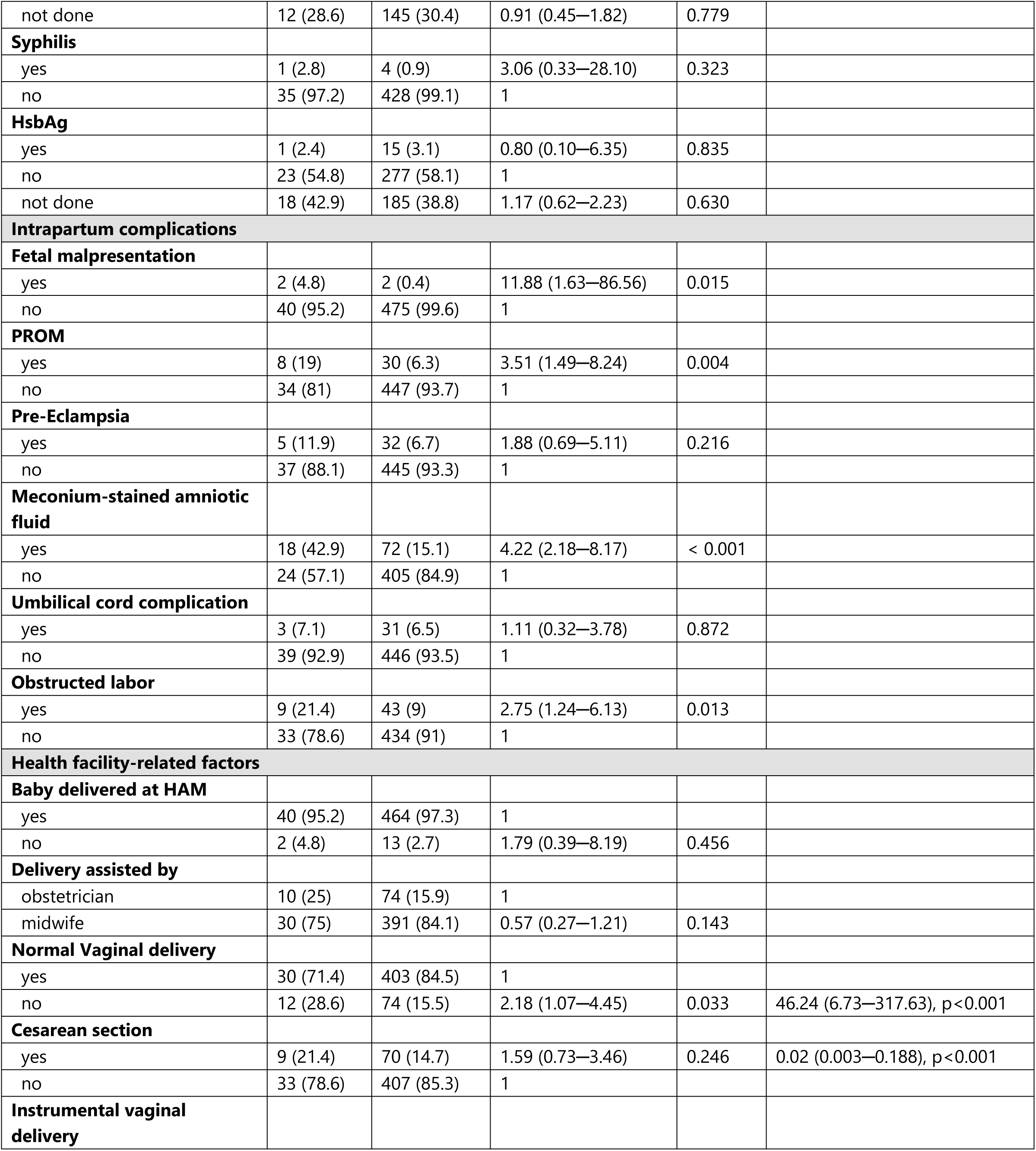
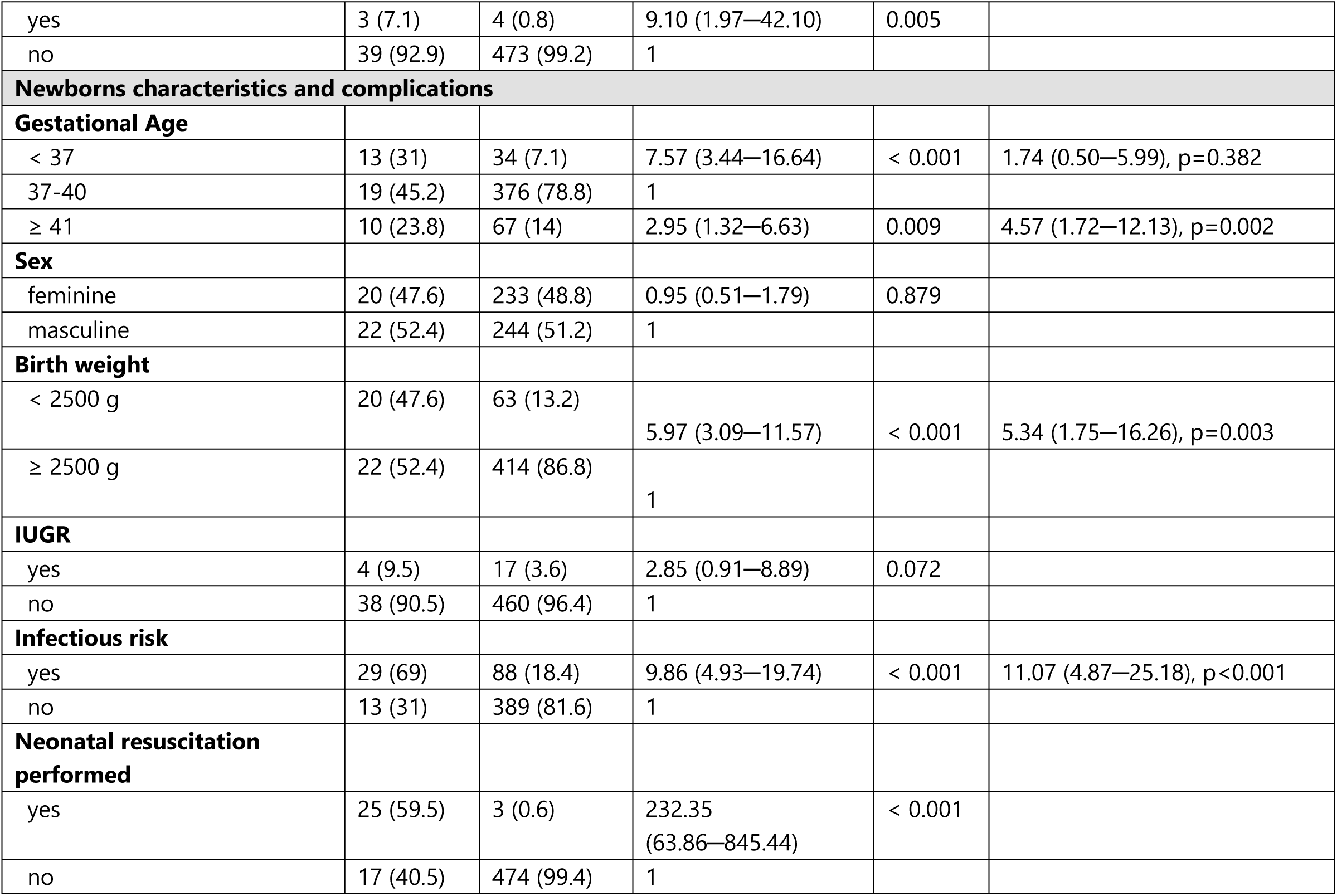
Factors associated with neonatal asphyxia among newborns delivered at Hospital Dr. Ayres de Menezes in Sao Tome & Principe - frequencies, percentages, CI for OR and aOR and p-value (univariable and multivariable analysis)

| Variables | asphyxia<br>n = 42<br>n (%) | no-asphyxia<br>n = 477<br>n (%) | cOR (95% CI) | p-value | aOR (95% CI) p-value |
| --- | --- | --- | --- | --- | --- |
| **Sociodemographic characteristics** |  |  |  |  |  |
| **Age** |  |  |  |  |  |
| 14-16 | 4 (9.5) | 25 (5.2) | 2.35 (0.75—7.39) | 0.142 |  |
| 17-19 | 7 (16.7) | 67 (14) | 1.54 (0.63—3.76) | 0.346 |  |
| 20-34 | 21 (50) | 309 (64.8) | 1 |  |  |
| ≥ 35 | 10 (23.8) | 76 (15.9) | 1.94 (0.88—4.28) | 0.103 |  |
| **Maternal education** |  |  |  |  |  |
| none | 1 (2.4) | 18 (3.8) | 0.54 (0.07—4.17) | 0.551 |  |
| primary | 28 (66.7) | 270 (56.6) | 1 |  |  |
| secondary | 8 (19) | 158 (33.1) | 0.49 (0.22—1.10) | 0.083 |  |
| higher | 5 (11.9) | 31 (6.5) | 1.56 (0.56—4.32) | 0.397 |  |
| **Maternal occupation** |  |  |  |  |  |
| housewife | 25 (59.5) | 301 (63.1) | 1 |  |  |
| student | 7 (16.7) | 29 (6.1) | 2.91 (1.16—7.30) | 0.023 | 5.30 (1.66—16.97), p=0.005 |
| employed | 10 (23.8) | 147 (30.8) | 0.82 (0.38—1.75) | 0.606 | 0.81 (0.33—1.97), p=0.645 |
| **Marital status** |  |  |  |  |  |
| union/married | 33 (78.6) | 377 (79) | 1 |  |  |
| single | 9 (21.4) | 100 (21) | 1.03 (0.48—2.22) | 0.944 |  |
| **Residence** |  |  |  |  |  |
| urban | 15 (38.5) | 216 (45.9) | 0.74 (0.38—1.44) | 0.374 |  |
| rural | 24 (61.5) | 255 (54.1) | 1 |  |  |
| **Improved water** |  |  |  |  |  |
| yes | 32 (76.2) | 345 (72.3) | 1 |  |  |
| no | 10 (23.8) | 132 (27.7) | 0.82 (0.39—1.71) | 0.591 |  |
| **Sanitation** |  |  |  |  |  |
| yes | 21 (50) | 251 (52.6) | 1 |  |  |
| open defecation | 21 (50) | 226 (47.4) | 1.11 (0.59—2.09) | 0.745 |  |
| **Preconception factors** |  |  |  |  |  |
| **Pregnancy planned** |  |  |  |  |  |
| yes | 6 (14.3) | 120 (25.2) | 1 |  |  |
| no | 27 (64.3) | 266 (55.8) | 2.03 (0.82—5.05) | 0.127 |  |
| missing | 9 (21.4) | 91 (19.1) | 1.98 (0.68—5.76) | 0.211 |  |
| **Ever use of family planning methods** |  |  |  |  |  |
| yes | 8 (24.2) | 98 (24.6) |  |  |  |
| no | 25 (75.8) | 300 (75.4) | 1.02 (0.45—2.34) | 0.961 |  |
| **Gravidity** |  |  |  |  |  |
| 1 | 12 (28.6) | 114 (23.9) | 1 |  |  |
| 2-4 | 18 (42.9) | 255 (53.5) | 0.67 (0.31—1.44) | 0.305 |  |
| ≥ 5 | 12 (28.6) | 108 (22.6) | 1.06 (0.46—2.45) | 0.900 |  |
| **Parity** |  |  |  |  |  |
| 0 | 15 (35.7) | 138 (28.9) | 1 |  |  |
| 1-4 | 21 (50) | 295 (61.8) | 0.66 (0.33—1.31) | 0.231 |  |
| ≥ 5 | 6 (14.3) | 44 (9.2) | 1.26 (0.46—3.43) | 0.659 |  |
| **Previous abortion** |  |  |  |  |  |
| yes | 16 (38.1) | 140 (29.4) | 1.48 (0.77—2.85) | 0.238 |  |
| no | 26 (61.9) | 337 (70.6) | 1 |  |  |
| **Previous stillbirth** |  |  |  |  |  |
| yes | 5 (11.9) | 48 (10.1) | 1.21 (0.45—3.22) | 0.706 |  |
| no | 37 (88.1) | 429 (89.9) | 1 |  |  |
| **Poor birth spacing** |  |  |  |  |  |
| yes | 9 (21.4) | 91 (19.1) | 1.16 (0.54—2.50) | 0.711 |  |
| no | 33 (78.6) | 386 (80.9) | 1 |  |  |
| **ANC** |  |  |  |  |  |
| **GA at first ANC visit** |  |  |  |  |  |
| ≤ 12 | 18 (48.6) | 254 (63.7) | 1 |  |  |
| > 12 | 19 (51.4) | 145 (36.3) | 1.85 (0.94—3.64) | 0.075 |  |
| **Number of ANC contacts** |  |  |  |  |  |
| 1-4 | 10 (25) | 65 (13.7) | 2.66 (1.08—6.55) | 0.034 |  |
| 5-7 | 19 (47.5) | 218 (46.1) | 1.51 (0.70—3.24) | 0.296 |  |
| ≥ 8 | 11 (27.5) | 190 (40.2) | 1 |  |  |
| **Obstetric ultrasound** |  |  |  |  |  |
| yes | 23 (54.8) | 284 (59.5) | 1 |  |  |
| no | 19 (45.2) | 193 (40.5) | 1.22 (0.64—2.29) | 0.546 |  |
| **Twin pregnancy** |  |  |  |  |  |
| yes | 8 (19) | 26 (5.5) | 4.08 (1.72—9.70) | 0.001 | 3.73 (1.21—11.47), p=0.022 |
| no | 34 (81) | 451 (94.5) | 1 |  |  |
| **Maternal anemia** |  |  |  |  |  |
| yes | 14 (33.3) | 147 (30.8) | 1.25 (0.60—2.61) | 0.554 |  |
| no | 17 (40.5) | 223 (46.8) | 1 |  |  |
| not done | 11 (26.2) | 107 (22.4) | 1.35 (0.61—2.98) | 0.460 |  |
| **Bacteriuria** |  |  |  |  |  |
| yes | 7 (16.7) | 148 (31) | 0.47 (0.19—1.14) | 0.096 |  |
| no | 19 (45.2) | 188 (39.4) | 1 |  |  |
| not done | 16 (38.1) | 141 (29.6) | 1.12 (0.56—2.26) | 0.746 |  |
| **Hyperglycemia** |  |  |  |  |  |
| yes | 1 (2.4) | 15 (3.1) | 0.73 (0.09—5.72) | 0.763 |  |
| no | 29 (69) | 317 (66.5) | 1 |  |  |
| not done | 12 (28.6) | 145 (30.4) | 0.91 (0.45—1.82) | 0.779 |  |
| **Syphilis** |  |  |  |  |  |
| yes | 1 (2.8) | 4 (0.9) | 3.06 (0.33—28.10) | 0.323 |  |
| no | 35 (97.2) | 428 (99.1) | 1 |  |  |
| **HsbAg** |  |  |  |  |  |
| yes | 1 (2.4) | 15 (3.1) | 0.80 (0.10—6.35) | 0.835 |  |
| no | 23 (54.8) | 277 (58.1) | 1 |  |  |
| not done | 18 (42.9) | 185 (38.8) | 1.17 (0.62—2.23) | 0.630 |  |
| **Intrapartum complications** |  |  |  |  |  |
| **Fetal malpresentation** |  |  |  |  |  |
| yes | 2 (4.8) | 2 (0.4) | 11.88 (1.63—86.56) | 0.015 |  |
| no | 40 (95.2) | 475 (99.6) | 1 |  |  |
| **PROM** |  |  |  |  |  |
| yes | 8 (19) | 30 (6.3) | 3.51 (1.49—8.24) | 0.004 |  |
| no | 34 (81) | 447 (93.7) | 1 |  |  |
| **Pre-Eclampsia** |  |  |  |  |  |
| yes | 5 (11.9) | 32 (6.7) | 1.88 (0.69—5.11) | 0.216 |  |
| no | 37 (88.1) | 445 (93.3) | 1 |  |  |
| **Meconium-stained amniotic fluid** |  |  |  |  |  |
| yes | 18 (42.9) | 72 (15.1) | 4.22 (2.18—8.17) | < 0.001 |  |
| no | 24 (57.1) | 405 (84.9) | 1 |  |  |
| **Umbilical cord complication** |  |  |  |  |  |
| yes | 3 (7.1) | 31 (6.5) | 1.11 (0.32—3.78) | 0.872 |  |
| no | 39 (92.9) | 446 (93.5) | 1 |  |  |
| **Obstructed labor** |  |  |  |  |  |
| yes | 9 (21.4) | 43 (9) | 2.75 (1.24—6.13) | 0.013 |  |
| no | 33 (78.6) | 434 (91) | 1 |  |  |
| **Health facility-related factors** |  |  |  |  |  |
| **Baby delivered at HAM** |  |  |  |  |  |
| yes | 40 (95.2) | 464 (97.3) | 1 |  |  |
| no | 2 (4.8) | 13 (2.7) | 1.79 (0.39—8.19) | 0.456 |  |
| **Delivery assisted by** |  |  |  |  |  |
| obstetrician | 10 (25) | 74 (15.9) | 1 |  |  |
| midwife | 30 (75) | 391 (84.1) | 0.57 (0.27—1.21) | 0.143 |  |
| **Normal Vaginal delivery** |  |  |  |  |  |
| yes | 30 (71.4) | 403 (84.5) | 1 |  |  |
| no | 12 (28.6) | 74 (15.5) | 2.18 (1.07—4.45) | 0.033 | 46.24 (6.73—317.63), p<0.001 |
| **Cesarean section** |  |  |  |  |  |
| yes | 9 (21.4) | 70 (14.7) | 1.59 (0.73—3.46) | 0.246 | 0.02 (0.003—0.188), p<0.001 |
| no | 33 (78.6) | 407 (85.3) | 1 |  |  |
| **Instrumental vaginal delivery** |  |  |  |  |  |
| yes | 3 (7.1) | 4 (0.8) | 9.10 (1.97–42.10) | 0.005 |  |
| no | 39 (92.9) | 473 (99.2) | 1 |  |  |
| **Newborns characteristics and complications** |  |  |  |  |  |
| **Gestational Age** |  |  |  |  |  |
| < 37 | 13 (31) | 34 (7.1) | 7.57 (3.44–16.64) | < 0.001 | 1.74 (0.50–5.99), p=0.382 |
| 37-40 | 19 (45.2) | 376 (78.8) | 1 |  |  |
| ≥ 41 | 10 (23.8) | 67 (14) | 2.95 (1.32–6.63) | 0.009 | 4.57 (1.72–12.13), p=0.002 |
| **Sex** |  |  |  |  |  |
| feminine | 20 (47.6) | 233 (48.8) | 0.95 (0.51–1.79) | 0.879 |  |
| masculine | 22 (52.4) | 244 (51.2) | 1 |  |  |
| **Birth weight** |  |  |  |  |  |
| < 2500 g | 20 (47.6) | 63 (13.2) | 5.97 (3.09–11.57) | < 0.001 | 5.34 (1.75–16.26), p=0.003 |
| ≥ 2500 g | 22 (52.4) | 414 (86.8) | 1 |  |  |
| **IUGR** |  |  |  |  |  |
| yes | 4 (9.5) | 17 (3.6) | 2.85 (0.91–8.89) | 0.072 |  |
| no | 38 (90.5) | 460 (96.4) | 1 |  |  |
| **Infectious risk** |  |  |  |  |  |
| yes | 29 (69) | 88 (18.4) | 9.86 (4.93–19.74) | < 0.001 | 11.07 (4.87–25.18), p<0.001 |
| no | 13 (31) | 389 (81.6) | 1 |  |  |
| **Neonatal resuscitation performed** |  |  |  |  |  |
| yes | 25 (59.5) | 3 (0.6) | 232.35 (63.86–845.44) | < 0.001 |  |
| no | 17 (40.5) | 474 (99.4) | 1 |  |  |

In the multivariable logistic analysis (Table 3), maternal occupation as student (aOR 5.30 95% CI 1.66─16.97; p=0.005), twin pregnancy (aOR 3.73 95% CI 1.21─11.47; p=0.022), delivery other than normal vaginal delivery (aOR 46.24 95% CI 6.73─317.63; p<0.001), gestational age greater or equal to 41 (aOR 4.57 95% CI 1.72─12.13; p=0.002), low birth weight (aOR 5.34 95% CI 1.75─16.26; p=0.003) and infectious risk (aOR 11.07 95% CI 4.87─25.18; p<0.001) were independently significantly associated with neonatal asphyxia. Cesarean section (aOR 0.02 95% CI 0.003─0.188; p<0.001) was found to be a protective factor.

## Discussion

The observed prevalence of birth asphyxia in this study was 8.1%, which is lower than rates reported in other low- and middle-income countries (LMICs) [23, 26, 43, 45, 50–52, 54].

For instance, a study in Ethiopia found a pooled prevalence of 22.52%, while another study reported rates of 18% in East African countries and 9.1% in the Central Africa region. [23, 43]. The lower incidence observed in STP may be attributable to differences in case definitions. In the present study, birth asphyxia was defined exclusively by a 5-minute APGAR score below 7. In contrast, other studies have employed broader criteria, including umbilical cord pH below 7, a 20-minute Apgar score below 7, evidence of multiorgan failure within 72 hours of life, or the occurrence of convulsions within the first 24 hours. [13, 23].

Neonatal asphyxia was independently associated with maternal occupation as a student and multiple gestation (maternal factors); intrapartum infectious risk and dystocic delivery (intrapartum factors); as well as post-term gestation, and low birth weight (neonatal factors).

In the present study, the odds of neonatal asphyxia were found to be approximately five times higher among mothers whose occupation was listed as “student.” This association may be attributable to the younger age and consequently lower educational attainment of these mothers, as well as their lack of stable employment [44–46]. These factors could limit their financial resources and access to adequate antenatal care, thereby increasing the risk of adverse neonatal outcomes.

Twin pregnancy has been identified as a significant risk factor for neonatal asphyxia, as multiple gestation is associated with a higher likelihood of obstetric and perinatal complications, such as preterm delivery, prolonged labor and low birth weight. In this study, twin pregnancy had approximately four times higher odds of neonatal asphyxia, which is also reported in other study conducted in Nepal [25]. This association reflects the added complexity of multiple gestation and underscores the importance of intensive monitoring and skilled obstetric care to prevent adverse outcomes, including birth asphyxia.

Infectious risk in our study was defined as the presence of maternal fever, prolonged rupture of membranes (PROM), and/or meconium-stained or foul-smelling amniotic fluid. Both premature and prolonged rupture of membranes have been consistently identified as factors associated with neonatal asphyxia, particularly in low-resource settings where timely diagnosis and intervention may be limited [50, 51]. PROM increases the risk of oligohydramnios, which can compromise umbilical cord blood flow and oxygen delivery to the fetus, and also facilitates the ascent of microorganisms, leading to intrauterine infection and subsequent neonatal sepsis [18, 21]. In our study, PROM was present in nearly one-fifth of asphyxia cases.

Meconium-stained amniotic fluid, another well-established risk factor for neonatal asphyxia [23–24, 45, 52–53] was also observed in a significant proportion of asphyxiated newborns in our cohort. The presence of meconium in the amniotic fluid is often a marker of fetal distress and, in low-resource settings, the lack of advanced monitoring and immediate intervention can increase the likelihood of meconium aspiration and hypoxic events [15, 53].

When considered collectively, neonates with any infectious risk had an elevenfold higher risk of developing neonatal asphyxia. This underscores the critical importance of early identification and management of maternal infections and PROM, especially in settings where access to rapid diagnostics, antibiotics, and emergency obstetric care may be limited. It is important to note that, due to data limitations, we were unable to assess the impact of maternal fever at the time of delivery in this study.

Relating to the mode of delivery, neonates with a dystocic delivery (both by cesarean section and instrumental vaginal delivery) had a 46 times higher risk of neonatal asphyxia which is consistent with the literature [23–24, 45, 50]. Interestingly, cesarean section emerged as a protective factor against neonatal asphyxia in our study (aOR 0.02; 95% CI 0.003–0.188; p<0.001). This finding may be partially explained by the relatively low number of asphyxia cases delivered via cesarean section (n=9), suggesting that timely surgical intervention in selected high-risk deliveries might have prevented intrapartum hypoxic events. However, this association should be interpreted with caution due to the small event count and potential confounding by indication.

Late-term (41+0–41+6 weeks) and post-term (>42+0 weeks) pregnancies are associated with increased risks of adverse perinatal outcomes, such as meconium-stained amniotic fluid, fetal distress, and perinatal mortality, all of which are more common in prolonged pregnancies [47–49]. In this study, births after 41 weeks of gestation had approximately five times higher odds of neonatal asphyxia. In low-resource settings like this one, limited access to timely obstetric interventions—such as labour induction, continuous fetal monitoring, and emergency cesarean section—can exacerbate the risks of neonatal asphyxia associated with prolonged pregnancies. Delays in recognizing and responding to fetal distress or placental insufficiency may lead to higher rates of asphyxia in these infants [17]. These systemic constraints make it challenging to prevent, detect, and manage complications that can lead to asphyxia, particularly in late-term and post-term births [15, 21].

Low birth weight (< 2500g) was also found to have five times higher odds of neonatal asphyxia, which is consistent with other studies where low birth weight is a well-identified risk factor associated with neonatal asphyxia [26, 54–56]. Low-birth-weight infants are often preterm, and may lack adequate surfactant, making them prone to respiratory distress and asphyxia [57]. Their limited brown fat stores also increase the risk of hypothermia, further worsening asphyxia [58]. Consequently, these neonates require prompt respiratory support, thermal care, and adequate nutrition to assist their transition to the extra-uterine environment. [23, 24]

Although preterm birth (<37 weeks of gestation) is widely recognized as a risk factor for neonatal asphyxia due to increased vulnerability to respiratory distress and physiological immaturity, in our study, this association was not statistically significant after adjustment for confounding factors (aOR 1.74; 95% CI 0.50–5.99; p=0.382). This suggests that the observed crude association may be mediated by related factors such as low birth weight or multiple gestation, which were independently associated with the outcome. In this setting, it is important to note that the risk of birth asphyxia among preterm infants may be exacerbated by the limited availability of timely antenatal corticosteroid administration, continuous intrapartum fetal monitoring, and advanced neonatal resuscitation — all of which are essential components of effective preterm birth management. [23, 45–46]

### Limitations

This study has several limitations that should be considered when interpreting the findings. First, the case definition relied solely on a 5-minute Apgar score <7, which, while widely used, may lack specificity in distinguishing the severity or underlying causes of neonatal asphyxia, especially in low-resource settings where confirmatory tests such as arterial blood gas analysis are not routinely available. Second, the retrospective reliance on clinical records may have introduced information bias, particularly for intrapartum and preconception variables. Third, due to the relatively small number of asphyxia cases (n=42), some associations—particularly those with wide confidence intervals—should be interpreted with caution. Lastly, residual confounding cannot be ruled out despite multivariable adjustment, given the complex interplay between maternal, obstetric, and neonatal factors in the pathophysiology of birth asphyxia.

## Conclusion

This study is the first to identify independent risk factors associated with neonatal asphyxia in Sao Tome and Principe. The findings highlight the multifactorial nature of the condition, involving maternal, intrapartum, and neonatal factors. Among maternal factors, student status and twin pregnancy were significantly associated with higher odds of neonatal asphyxia, suggesting the influence of socioeconomic vulnerability and obstetric complexity. Intrapartum contributors included the presence of infectious risk and dystocic deliveries, emphasizing the importance of timely recognition and management of labor complications. Neonatal factors such as post-term birth and low birth weight were also independently associated with increased risk, reflecting the need for enhanced foetal surveillance and delivery planning.

These results underscore the urgent need for integrated strategies aimed at strengthening antenatal risk identification, improving labour management, and ensuring readiness for neonatal resuscitation. In resource-limited settings like Sao Tome and Principe, even incremental improvements in maternal and newborn care—particularly among vulnerable subgroups—could have a meaningful impact on reducing the incidence and consequences of neonatal asphyxia.

## Availability of data and materials

All data generated or analysed during this study are included in this paper or in other published article [8–14].

## Abbreviations

HAM: Hospital Dr. Ayres de Menezes
ANC: antenatal care
WHO: World Health Organization
NTD: Neglected Tropical Diseases.

## Data Availability

All relevant data are within the manuscript and its Supporting Information files

## Acknowledgements

A special remark for the late Professor João Luís Baptista PhD MD - AV research cosupervisor - a great man who was a thinker and a fighter for Sao Tome and Príncipe improvement of public health. We are indebted to all the women who participated in the study. The authors would like to thank Elizabeth Carvalho and the 1) medical team and nurses of Hospital Ayres de Menezes Maternity for their support, especially the chief-nurse Paulina Oliveira. We would like to acknowledge Instituto Camões, I.P. for logistic support in Sao Tome & Principe.

## Authors’ contributions

AV and FP conceived and designed the study. AV was responsible for field activities and data collection. CB, MV, VC and AV drafted the manuscript. AV and FP critically reviewed the study, provided ongoing input throughout its development, and contributed to the manuscript revision. BC performed the statistical analysis and participated in the manuscript review. SS and NB contributed to the study design at the country level. All authors read and approved the final version of the manuscript.

## Supporting information

## References

1. World Health Organization. Guidelines on basic newborn resuscitation, 2012. Available at: https://www.who.int/teams/maternal-newborn-child-adolescent-health-and-ageing/newborn-health/perinatal-asphyxia

2. Rainaldi MA, Perlman JM. Pathophysiology of birth asphyxia. Clin Perinatol. (2016) 43:409–22. doi: 10.1016/j.clp.2016.04.002

3. Gillam-Krakauer M, Gowen Jr CW. Birth Asphyxia. [Updated 2023 Aug 14]. In: StatPearls [Internet]. Treasure Island (FL): StatPearls Publishing; 2024 Jan-. Available from: https://www.ncbi.nlm.nih.gov/books/NBK430782/

4. American Academy of Pediatrics Committee on Fetus and Newborn; American College of Obstetricians and Gynecologists Committee on Obstetric Practice. The Apgar Score. Pediatrics. 2015;136(4):819–822. doi:10.1542/peds.2015-2651

5. Country Office Annual Report 2023, Sao Tome and Principe, UNICEF. Available at: https://www.unicef.org/media/152996/file/Sao-Tome-and-Principe-2023-COAR.pdf

6. Key demographic indicators in Sao Tome and Principe, Country profiles, UNICEF. Available at: https://data.unicef.org/country/stp/

7. Sao Tome and Principe — Causes of Neonatal Mortality, 2016 by WHO-MCEE estimates for child causes of death, 2000-2016 in Maternal and Newborn Health Disparities in São Tomé and Príncipe, UNICEF

8. Vasconcelos A, Bandeira N, Sousa S, Pereira F, Machado MC. Adolescent pregnancy in Sao Tome and Principe: a cross-sectional hospital-based study. BMC Pregnancy Childbirth. 2022;22(1):332. Available from: 10.1186/s12884-022-04632-z

9. Vasconcelos A, Bandeira N, Sousa S, Machado MC, Pereira F. Adolescent pregnancy in Sao Tome and Principe: are there different obstetric and perinatal outcomes? BMC Pregnancy Childbirth. 2022;22(1):453. Available from: 10.1186/s12884-022-04779-9

10. Vasconcelos A, Sousa S, Bandeira N, Alves M, Papoila AL, Pereira F, Machado MC. Intestinal parasitic infections, treatment and associated factors among pregnant women in Sao Tome and Principe: A cross-sectional study. J Trop Med. 2022;2022:7492020. Available from: 10.1155/2022/7492020

11. Vasconcelos A, Sousa S, Bandeira N, Alves M, Papoila AL, Pereira F, Machado MC. Antenatal screenings and maternal diagnosis among pregnant women in Sao Tome & Principe-Missed opportunities to improve neonatal health: A hospital-based study. PLOS Global Public Health. 2022;2. Available from: 10.1371/journal.pgph.0001444

12. Vasconcelos A, Sousa S, Bandeira N, Alves M, Papoila AL, Pereira F, Machado MC. Determinants of antenatal care utilization–contacts and screenings–in Sao Tome & Principe: a hospital-based cross-sectional study. Archives of Public Health.2023; 81(1), 107. Available from: 10.1186/s13690-023-01123-1

13. Vasconcelos A, Sousa S, Bandeira N, Alves M, Papoila A, Pereira F, et al. (2023) Adverse birth outcomes and associated factors among newborns delivered in Sao Tome & Principe: A case–control study. PLoS ONE 18(7): e0276348. 10.1371/journal.pone.0276348

14. Vasconcelos A, Sousa S, Bandeira N, Alves M, Papoila AL, Pereira F and Machado MC (2024) Factors associated with perinatal and neonatal deaths in Sao Tome & Principe: a prospective cohort study. Front. Pediatr. 12:1335926. doi: 10.3389/fped.2024.1335926

15. Lawn JE, Blencowe H, Oza S, et al. (2014). Every Newborn: progress, priorities, and potential beyond survival. Lancet, 384(9938), 189–205.

16. Lee AC, Cousens S, Wall SN, et al. (2013). Neonatal resuscitation and immediate newborn assessment and stimulation for the prevention of neonatal deaths: a systematic review, meta-analysis and Delphi estimation of mortality effect. BMC Public Health, 13(Suppl 3), S12.

17. Goldenberg RL, McClure EM. (2012). Maternal, fetal and neonatal mortality: lessons learned from historical changes in high-income countries and their potential application to low-income countries. Matern Health Neonatol Perinatol, 1(1), 3.

18. Goldenberg RL, McClure EM, Saleem S, Reddy UM. (2010). Infection-related stillbirths. Lancet, 375(9724), 1482–1490.

19. Newton O, English M. (2010). Newborn resuscitation: defining best practice for low-income settings. Trans R Soc Trop Med Hyg, 104(4), 261–268.

20. Lawn, J. E., Kerber, K., Enweronu-Laryea, C., & Massee Bateman, O. (2009). Newborn survival in low resource settings--are we delivering?. BJOG : an international journal of obstetrics and gynaecology, 116 *Suppl 1*, 49–59. 10.1111/j.1471-0528.2009.02328.x

21. World Health Organization. (2016). Standards for improving quality of maternal and newborn care in health facilities.

22. UNICEF. (2021). Levels and Trends in Child Mortality: Report 2021.

23. Bayih WA, Birhane BM, Belay DM, et al. The state of birth asphyxia in Ethiopia: An umbrella review of systematic review and meta-analysis reports, 2020. Heliyon. 2021; 7(10):e08128. 10.1016/j.heliyon.2021.e08128 PMID: 34746456

24. Wosenu L, Worku AG, Teshome DF, Gelagay AA. Determinants of birth asphyxia among live birth newborns in University of Gondar referral hospital, northwest Ethiopia: A case-control study. PLoS One. 2018 Sep 7;13(9):e0203763. doi: 10.1371/journal.pone.0203763. PMID: 30192884; PMCID: PMC6128623.

25. Lee AC, Mullany LC, Tielsch JM, Katz J, Khatry SK, LeClerq SC, Adhikari RK, Shrestha SR, Darmstadt GL. Risk factors for neonatal mortality due to birth asphyxia in southern Nepal: a prospective, community-based cohort study. Pediatrics. 2008 May;121(5):e1381–90. doi: 10.1542/peds.2007-1966. PMID: 18450881; PMCID: PMC2377391.

26. Ayebare, E., Hanson, C., Nankunda, J. et al. Factors associated with birth asphyxia among term singleton births at two referral hospitals in Northern Uganda: a cross sectional study. BMC Pregnancy Childbirth 22, 767 (2022). 10.1186/s12884-022-05095-y

27. INE e UNICEF. 2020. Inquérito de Indicadores Múltiplos 2019, Relatório final. São Tomé, São Tomé e Príncipe: Instituto Nacional de Estatística e Fundo das Nações Unidas para a Infância. Available at: https://washdata.org/sites/default/files/2022-02/Sao%20Tome%20e%20Principe%202019%20MICS-po.pdf

28. Lwanga SK, Lemeshow S. Sample size determination in health studies: a practical manual. 1991.

29. Sample size calculator by raosoft, inc. In: Raosoft.com [Internet]. [cited 8 May 2023]. Available: http://www.raosoft.com/samplesize.html.

30. Stadther D, Meyer CL. Sample size software. In: Ncss.com [Internet]. 10 Aug 2012 [cited 8 May 2023]. Available: https://www.ncss.com/software/pass/.

31. Abadiga M, Mosisa G, Tsegaye R, et al. Determinants of adverse birth outcomes among women delivered in public hospitals of Ethiopia, 2020. Archives of Public Health. 2022; 80(1):1–7. 10.1186/s13690-021-00776-0.

32. Sao Tome and Principe WHO statistical profile. WHO Libr. 2015. https://www.who.int/gho/countries/stp.

33. Pimentel J, Ansari U, Omer K, Gidado Y, Baba MC, Andersson N, et al. Factors associated with short birth interval in low- and middle-income countries: a systematic review. BMC Pregnancy Childbirth. (2020) 20:156. doi: 10.1186/s12884-020-2852-z

34. Mulowooza J, Santos N, Isabirye N, Inhensiko I, Sloan NL, Shah S, et al. Midwife-performed checklist and ultrasound to identify obstetric conditions at labour triage in Uganda: a quasi-experimental study. Midwifery. (2021) 96:102949. doi: 10.1016/j.midw.2021.102949

35. Chiabi A, Takou V, Mah E, Nguefack S, Siyou H, Takou V, et al. Risk factors for neonatal mortality at the yaounde gynaeco-obstetric and pediatric hospital. Cameroon.Iran J Pediatr. (2014) 24(4):393–400.

36. Tewabe T, Mohammed S, Tilahun Y, Melaku B, Fenta M, Dagnaw T, et al. Clinical outcome and risk factors of neonatal sepsis among neonates in felege hiwot referral hospital, bahir dar, amhara regional state, North West Ethiopia 2016: a retrospective chart review. BMC Res Notes. (2017) 10:265. doi: 10.1186/s13104-017-2573-1

37. Shaikh S, Shaikh AH, Shaikh SA, Isran B. Frequency of obstructed labor in teenage pregnancy. Nepal J Obstet Gynaecol. (2012) 7(1):37–40. doi: 10.3126/njog.v7i1.8834

38. Yang J, Wang M, Tobias DK, et al. Dietary diversity and diet quality with gestational weight gain and adverse birth outcomes, results from a prospective pregnancy cohort study in urban Tanzania. Maternal & Child Nutrition. 2022; 18(2):e13300. 10.1111/mcn.13300 PMID: 34908233

39. Chawanpaiboon S, Vogel JP, Moller A-B, Lumbiganon P, Petzold M, Hogan D, et al. Global, regional, and national estimates of levels of preterm birth in 2014: a systematic review and modelling analysis. The Lancet global health. 2019; 7(1):e37—e46. 10.1016/S2214-109X(18)30451-0 PMID: 30389451.

40. Mandy GT. Preterm birth: Definitions of prematurity, epidemiology, and risk factors for infant mortality. UpToDate Retrieved September2024 https://www.uptodate.com/contents/preterm-birth-definitions-of-prematurity-epidemiology-and-risk-factors-for-infant-mortality

41. Huynh B, Kermorvant-Duchemin E, Herindrainy P et al. Bacterial Infections in Neonates, Madagascar, 2012–2014. Emerging Infectious Diseases. 2018; 24(4), 710–717. 10.3201/eid2404.161977. PMID: 29553312

42. Accrombessi M, Zeitlin J, Massougbodji A, Cot M, Briand V. What do we know about risk factors for fetal growth restriction in Africa at the time of sustainable development goals? A scoping review. Paediatr Perinat Epidemiol. (2018) 32 (2):184–96. doi: 10.1111/ppe.12433

43. Workineh Y, Semachew A, Ayalew E, Animaw W, Tirfie M, Birhanu M. Prevalence of perinatal asphyxia in East and Central Africa: systematic review and meta-analysis. Heliyon. 2020 Apr 1; 6(4):e03793. 10.1016/j.heliyon.2020.e03793 PMID: 32368646

44. Igboanugo S, Chen A, Mielke JG. Maternal risk factors for birth asphyxia in low-resource communities. A systematic review of the literature. J Obstet Gynaecol. 2020 Nov;40(8):1039–1055. doi: 10.1080/01443615.2019.1679737. Epub 2019 Dec 11. PMID: 31825270.

45. Desalew, et al., International Journal of Health Sciences. Determinants of Birth Asphyxia Among Newborns in Ethiopia: A Systematic Review and Meta-Analysis, 2020.

46. Ikechebelu JI, Eleje GU, Onubogu CU, et al. Incidence, predictors and immediate neonatal outcomes of birth asphyxia in Nigeria. BJOG. 2024;131 Suppl 3:88–100. doi:10.1111/1471-0528.17816

47. Hovi M, Raatikainen K, Heiskanen N, Heinonen S. Obstetric outcome in post-term pregnancies: time for reappraisal in clinical management. Acta Obstet Gynecol Scand. 2006;85(7):805–809. doi:10.1080/00016340500442472

48. Patel TL, Rathod DA. Study of maternal and perinatal outcome in postdated pregnancy.Int J Reprod Contracept Obstet Gynecol 2022;11:1507–11.

49. Maoz O, Wainstock T, Sheiner E, Walfisch A. Immediate perinatal outcomes of postterm deliveries. J Matern Fetal Neonatal Med. 2019;32(11):1847–1852. doi:10.1080/14767058.2017.1420773

50. Msisiri LS, Kibusi SM, Kimaro FD. Risk Factors for Birth Asphyxia in Hospital-Delivered Newborns in Dodoma, Tanzania: A Case-Control Study. SAGE Open Nursing. 2024;10. doi:10.1177/23779608241246874

51. Wodajo L. T., Bedie N. A., Mengesha S. T. (2019). Magnitude and determinants of birth asphyxia : Unmatched case control study Assela referral teaching hospital, Arsi Zone, Ethiopia. Global Journal of Reproductive Medicine, 7(1), 21–29. 10.19080/GJORM.2019.07.555705

52. Tegegnework, S.S., Gebre, Y.T., Ahmed, S.M., et al. Determinants of birth asphyxia among newborns in Debre Berhan referral hospital, Debre Berhan, Ethiopia: a case-control study. BMC Pediatr 22, 165 (2022). 10.1186/s12887-022-03223-3

53. Mecacci F, et al. (2015). Late-term and post-term pregnancies: Obstetric management and perinatal outcomes. Minerva Ginecol, 67(5), 453–462.

54. Tasew, H., Zemicheal, M., Teklay, G. et al. Risk factors of birth asphyxia among newborns in public hospitals of Central Zone, Tigray, Ethiopia 2018. BMC Res Notes 11, 496 (2018). 10.1186/s13104-018-3611-3

55. Alemu AY, Belay GM, Berhanu M, Minuye B. Determinants of neonatal mortality at neonatal intensive care unit in Northeast Ethiopia: unmatched case-control study. Trop Med Health. (2020) 48(1):1–0. doi:10.1186/s41182-019-0188-z

56. Techane MA, Alemu TG, Wubneh CA, et al. The effect of gestational age, low birth weight and parity on birth asphyxia among neonates in sub-Saharan Africa: systematic review and meta-analysis: 2021. Ital J Pediatr. 2022;48(1):114. Published 2022 Jul 15. doi:10.1186/s13052-022-01307-5

57. Yadav S, Lee B. Neonatal Respiratory Distress Syndrome. [Updated 2023 Jul 25]. In: StatPearls [Internet]. Treasure Island (FL): StatPearls Publishing; 2025 Jan-. Available from: https://www.ncbi.nlm.nih.gov/books/NBK560779/

58. Kumar, V., Shearer, J. C., Kumar, A., & Darmstadt, G. L. (2009). Neonatal hypothermia in low resource settings: a review. Journal of perinatology: official journal of the California Perinatal Association, 29(6), 401–412. 10.1038/jp.2008.233

